# Cardiovascular Risks Among the Transgender Population in Rural United States: Spotlight on the Appalachian Region

**DOI:** 10.1101/2023.09.13.23295518

**Authors:** Ebubechukwu Ezeh, Maddie Perdoncin, Chukwuemeka Ogbu, Esiemoghie Akhigbe, Mohammed Al-Hiari, Archana Ramalingam, Elizabeth Saunders, Jason Mader, Patricia F. Rodriguez Lozano

## Abstract

**Background:** Gender diverse populations have disproportionately worse cardiovascular (CV) outcomes. However, the data on the prevalence of cardiovascular risk factors among the transgender population in rural Appalachia remains limited. The purpose of this study was to determine the prevalence of CV risk factors among the transgender population in rural Appalachia.

**Methods:** This retrospective case-control study from an Appalachian university teaching hospital clinic matched transgender individuals with controls from the same clinic. Logistic regression model was performed to determine the prevalence of CV risk factors among the transgender population in comparison to their cisgender counterparts.

**Results:** The total number of transgender and cisgender individuals identified were 89 and 69, respectively. After adjusting for age and family history of atherosclerotic CV disease, transgender status remained independently associated with tobacco use (adjusted odds ratio [aOR] 6.64 [2.59-17.01]) and prediabetes/diabetes mellitus (aOR 3.98 [1.05-15.15]). Among the transgender population, there were increased odds of obesity (aOR 13.39 [3.30-54.30] and hyperlipidemia (aOR 3.46 [1.03-11.59]) in the transgender male group compared to the transgender female group. In another subgroup analysis, transgender individuals who underwent surgical and/or hormonal treatment had significantly higher odds of tobacco use (aOR 6.67 [2.80-15.98]), statin need (aOR 3.97 [1.14-13.81], and alcohol use (aOR 11.31 [1.34-95.53]). Increased statin use tends to be associated with normal LDL levels.

**Conclusions:** In rural Appalachia, transgender status is associated with disproportionately higher odds of tobacco use, elevated blood glucose and other CV risk factors.

## Introduction

Cardiovascular diseases (CVD) are the leading cause of mortality globally (1). Between 1990 and 2019, the number of deaths due to CVD has climbed from 12.1 million to 18.6 million worldwide.^1^ Identifying and minimizing potentially modifiable risk factors remains crucial in preventing these outcomes. In recent years, there has been a growing recognition of the importance of understanding the health disparities faced by minority populations, including the transgender community, in relation to CVD. While cisgender is used to describe people whose gender is congruent with their birth sex, the term “transgender” describes individuals whose inner sense of gender identity differs from their sex assigned at birth.^2^

The prevalence of individuals identifying as transgender has significantly increased in the past decade, with national data from 2021 estimating that over 2 million adults, or greater than 1% of the United States population, currently identify as transgender.^3^ This rise in the transgender population has been accompanied by a disproportionate incidence of cardiovascular mortality and morbidity within the community, as evidenced by long-term outcome studies.^4^ A self-reported surveillance study discovered that transgender men and women experienced a significantly increased prevalence of myocardial infarction compared to their cisgender counterparts.^5^

A recent study shows that West Virginia has the highest per capita rate of transgender youth in the country at just over 1%.^6^ As sociocultural acceptance continues to evolve, more transgender individuals are expected to seek care from healthcare providers. This trend is particularly important in Appalachian states, where concerns regarding limited access to healthcare are on the rise. Further, there is a disproportionate burden of CVD and associated risk factors in the Appalachian region.^7^ One study found that obesity and diabetes mellitus, common risk factors in central Appalachia, more than doubled the odds of having hypertension.^7^

Gender-affirming hormone therapy (GAHT) has also increased, aiming to align physical appearances and gender expression with preferred gender identity, which helps relieve gender dysphoria.^8^ Although GAHT is essential for the quality of life of transgender patients, the risk of developing CVD remains high. Understanding the potential risks of such treatment among this population is crucial to make informed decisions regarding GAHT.

Multiple recent studies have reported significantly higher rates of CVD risk factors among transgender patients receiving GAHT.^8–11^ A recent scientific statement by the American Heart Association concluded that the higher rates of CVD and death among TGD individuals, while related to traditional risk factors, are exacerbated by chronically high levels of stress driven by a lifetime of discrimination, the threat of violence, lack of affordable housing and access to health care.^12^ Despite improvements in cardiovascular health, there has been a concomitant increase in cardiometabolic risk factors within the transgender population^10^. This maybe related to the use of GAHT, but health research on this topic remains limited, with discrepant results.

In this retrospective case-control study, we aim to evaluate the prevalence and odds of CVD risk factors in the transgender population in our rural Appalachian community and propose a comprehensive approach to managing this susceptible population.

## Methods

### Study Design and Participants

We conducted a retrospective observational study using data from 92 self-identified transgender individuals and 73 randomly selected and matched cisgender individuals. Participants were recruited from a tertiary care center in West Virginia using a model similar to Denby et al.^13^ We selected individuals who identify as transgender and have had at least two visits to our physician-run adult transgender clinic. We also recruited cisgender individuals who visited our regular clinic as the control group. Individuals less than 18 years of age and those with a diagnosis of HIV were excluded. The Marshall University Institutional Review Boards and Ethics Review Board approved the study.

### Data Collection and Measures

We extracted baseline demographics, past medical history, and laboratory studies from participants’ electronic medical records. Age was recorded in years, and biological sex was categorized as male or female. The history of aspirin and statin use, obesity (BMI > 30 kg/m^2^), tobacco use, family history of coronary artery disease (CAD), and alcohol use (defined as >1 drink per day) were ascertained and categorized accordingly.

Laboratory values for low-density lipoprotein (LDL) and glycated hemoglobin (HbA1c) were collected within two months of the initial visit. LDL levels greater than 100 were considered high. The history of diabetes mellitus (DM) diagnosis and medication use was ascertained for observations with missing HbA1C values in the medical chart. A composite variable was created using HbA1C levels and medical records of DM, categorized as either no DM/normal HbA1C or DM and/or HbA1c > 5.7%. An HbA1c level of >5.7% signifies that the person has a higher-than-normal blood glucose level. This value may indicate an increased risk for diabetes or prediabetes. According to guidelines [12], hypertension was defined based on a blood pressure reading: blood pressure > 160/90 mm Hg at one visit or > 140/90 mm Hg at two subsequent visits.

### Statistical Analysis

Bivariate analyses were conducted using the Rao-Chi-Square test for categorical risk factors and the Student’s t-test for continuous variables. Covariates were selected based on prior literature review and then statistically selected through backward selection. Sensitivity analysis performed multiple imputations of missing data. We reported the results of the listwise estimates.

The dependent variables (CV risk factors) were measured dichotomously. We estimated unadjusted and adjusted binary logistic regression models for having these risk factors in the cases and controls (transgender vs. cisgender). Logistic regression models were also conducted in both cases and controls using surgery/hormone treatment as a predictor. Finally, logistic regression was performed among cases with transgender sex (transgender male vs transgender female) as a predictor. We tabulated the results of logistic regression models as odds ratios (ORs) and 95% confidence intervals (CIs) and adjusted for age and family history of atherosclerotic cardiovascular disease (ASCVD) in all models.

Multicollinearity between independent variables was assessed using tolerance estimates and variance inflation factors. The statistical hypothesis was tested using two-sided p < 0.05 as the significance level or by non-overlapping 95% confidence intervals. Analyses were performed in SAS (version 9.4, Cary NC) and graphing in R (R Core Team, 2020) and Microsoft Excel (2016 version).

## Results

Our study assessed the association between CV risk factors and transgender status while accounting for surgery and hormone therapy. While a transgender male is someone who identifies as a male but was assigned female sex at birth, a transgender female is someone who identifies as female but was assigned male sex at birth. A cisgender male is someone who identifies as a male and was assigned male sex at birth while a cisgender female was assigned female sex at birth and identifies as a female. The study population comprised 158 individuals aged 18-57 years, with an average age of 29.2 years for the transgender group and 29.9 years for the cisgender group.

Mean LDL levels were 105.8 mg/dL (±37.5) across all the transgender groups and 108.2 mg/dL (±31.3) across all cisgender groups (p value 0.7223) (figure 1). Baseline participant characteristics are presented in figure 2. In our study, transgender males constituted 63% of the total transgender population, while the cisgender group consisted of 26% males. Transgender individuals had a higher prevalence of HTN (16.7% vs. 15.9%), tobacco use (41.0% vs. 11.6%), alcohol consumption (10.3% vs. 1.5%), prediabetes/DM (DM and/or HbA1C > 5.7%; 16.5% vs. 4.4%), and statin use (15.8% vs. 5.8%) than their cisgender counterparts. In contrast, cisgender participants had a higher prevalence of aspirin use (2.9% vs. 2.6%), high LDL levels (66.7% vs. 55.2%), and a family history of ASCVD (29.7% vs. 15.8%). The prevalence of obesity was similar between the cisgender and transgender participants (52.5% vs 53.6%).

**Figure 1:**
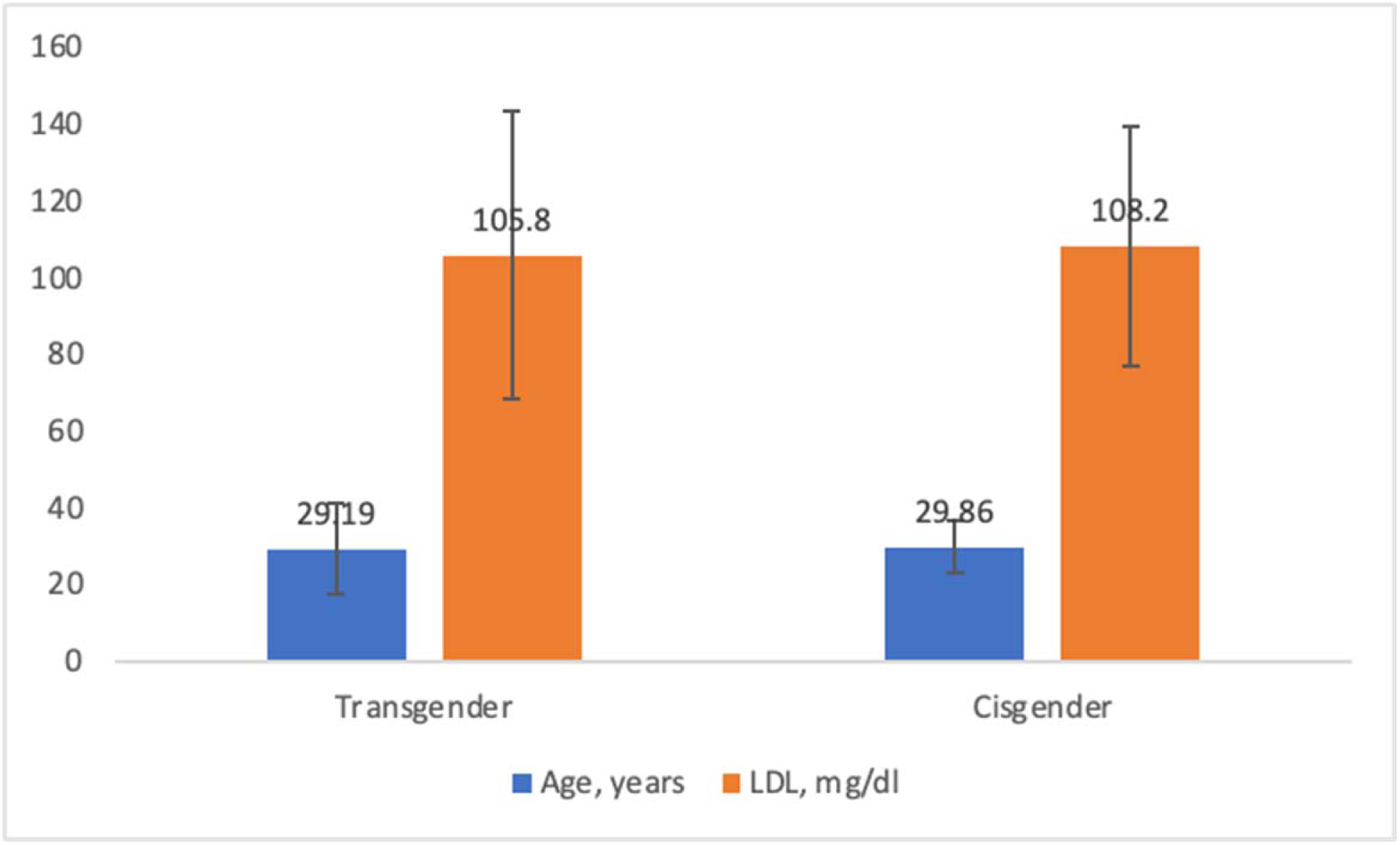
Mean age (years) and LDL (mg/dl) distribution in the transgender and cisgender populations.

**Figure 2:**
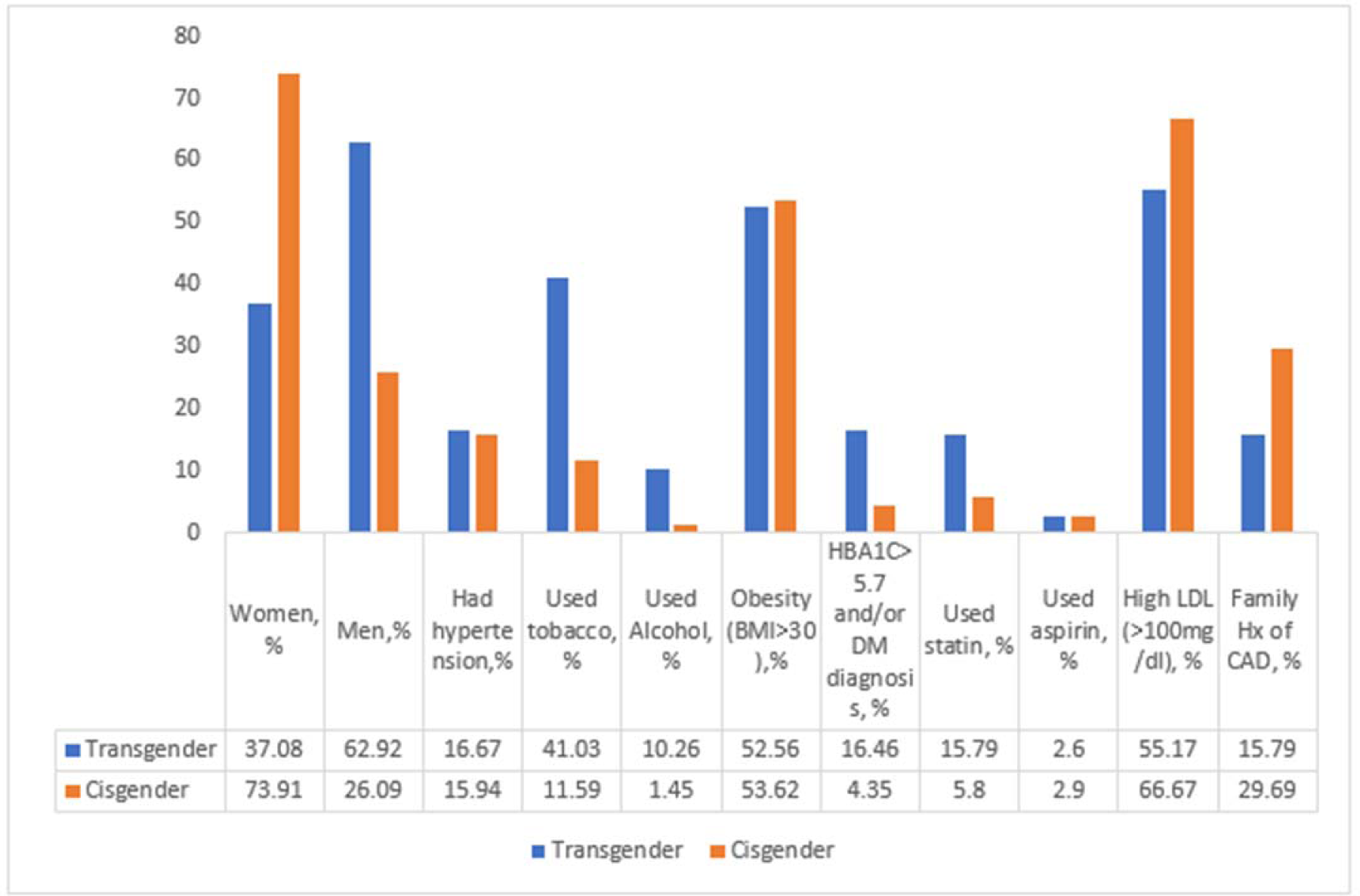
Percent prevalence of cardiovascular risk factors in the transgender and cisgender populations.

The adjusted logistic regression analysis of cardiovascular risk factors among transgender and cisgender populations are shown in figure 3. After adjusting for age and family history of ASCVD, transgender individuals exhibited a more than 6-fold increase in tobacco use (aOR, 6.64 [95% CI, 2.59-17.01]) and a 4-fold increase in prediabetes/diabetes (aOR, 3.98 [95% CI, 1.05-15.15]) compared to cisgender individuals. Although not statistically significant, there were increased odds of statin requirement, obesity, and alcohol use among the transgender population compared to the cisgender population (p < 0.05).

**Figure 3:**
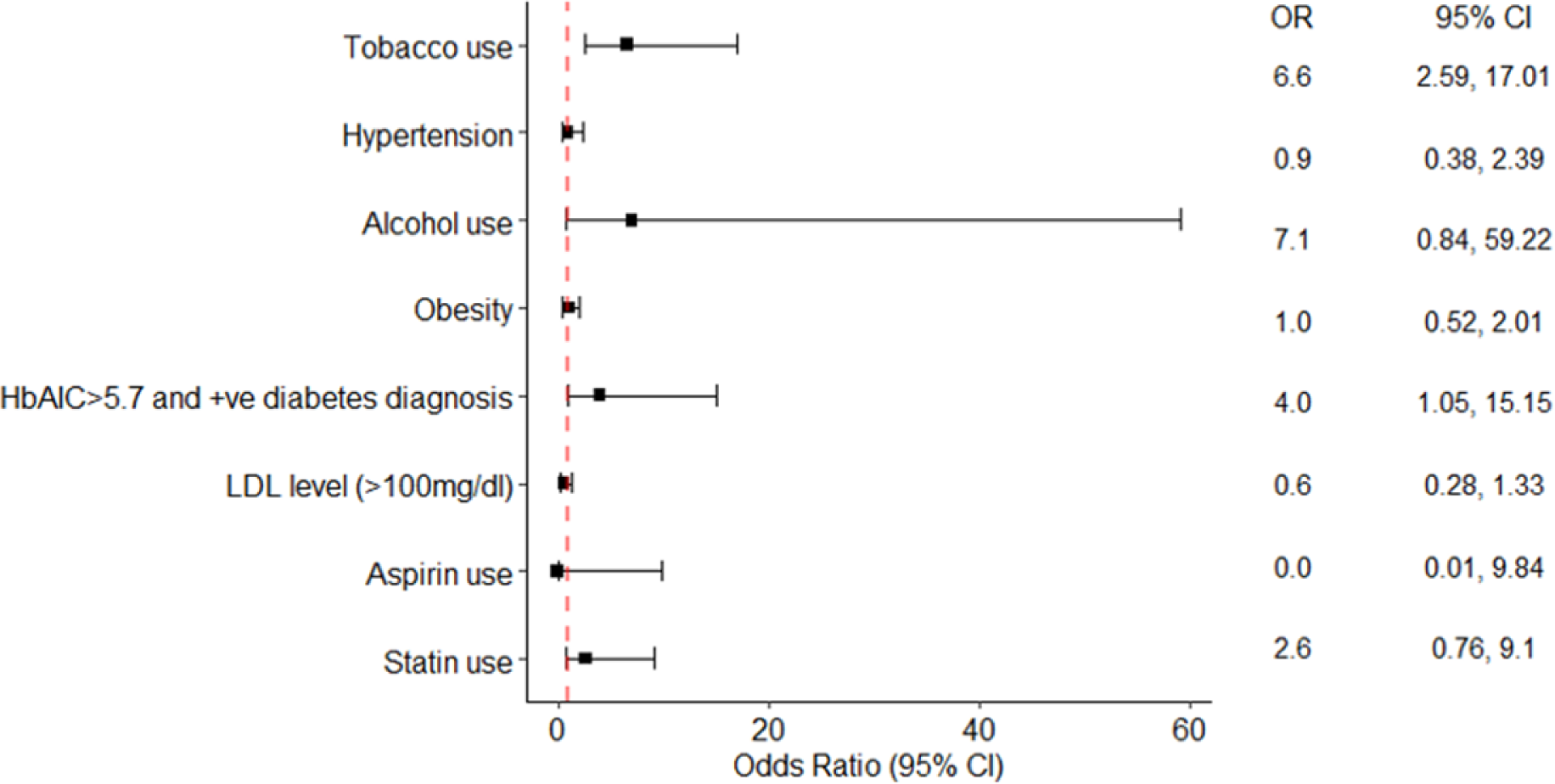
Adjusted multivariate logistic regression of cardiovascular risk factors among cases and controls.

Cardiovascular risk factors among transgender subgroups are presented in Table 1. Transgender males had significantly higher odds of obesity (unadjusted OR 4.85(1.77-13.29), p value 0.002; aOR, 13.39 [95% CI, 3.30-54.30], p value 0.0083) and hyperlipidemia (unadjusted OR 3.00(1-9.01), p value 0.052) aOR, 3.46 [95% CI, 1.03-11.59], p value 0.045) compared to transgender females.

**Table 1:**
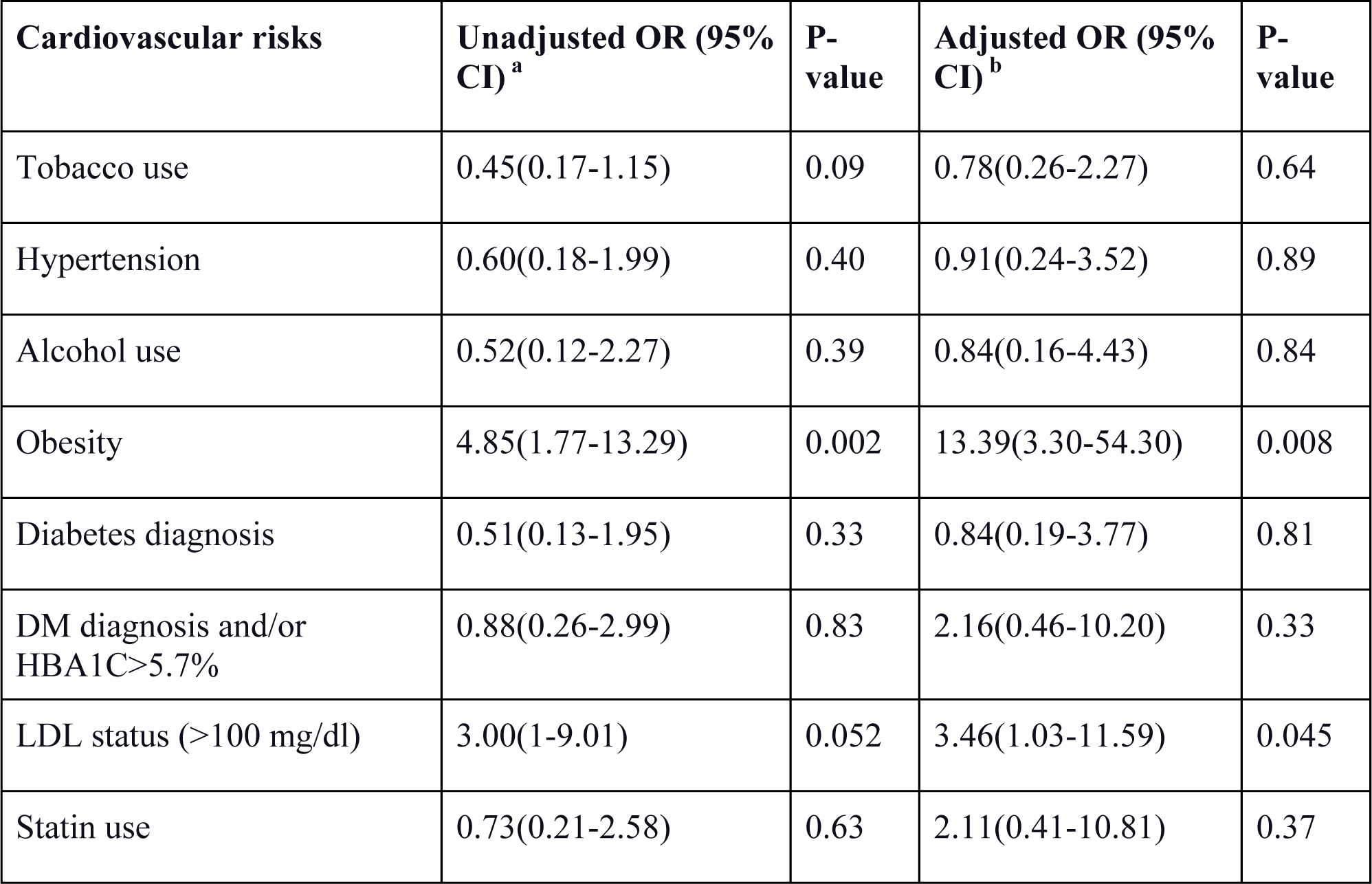
Unadjusted and adjusted multivariate logistic regression of cardiovascular risk factors among transgender males vs transgender females. CI, confidence interval; DM, Diabetes Mellitus; HbA1C, glycated hemoglobin; LDL, Low-Density Lipoprotein; ^a^ Cardiac risk factor as the dependent variable in regression models and transgender status (FTM vs. MTF) as a predictor in the model. ^b^ Each model adjusted for the age and family history of ASCVD.

| **Cardiovascular risks** | **Unadjusted OR (95% CI)<sup>a</sup>** | **P-value** | **Adjusted OR (95% CI)<sup>b</sup>** | **P-value** |
| --- | --- | --- | --- | --- |
| Tobacco use | 0.45(0.17-1.15) | 0.09 | 0.78(0.26-2.27) | 0.64 |
| Hypertension | 0.60(0.18-1.99) | 0.40 | 0.91(0.24-3.52) | 0.89 |
| Alcohol use | 0.52(0.12-2.27) | 0.39 | 0.84(0.16-4.43) | 0.84 |
| Obesity | 4.85(1.77-13.29) | 0.002 | 13.39(3.30-54.30) | 0.008 |
| Diabetes diagnosis | 0.51(0.13-1.95) | 0.33 | 0.84(0.19-3.77) | 0.81 |
| DM diagnosis and/or HbA1C>5.7% | 0.88(0.26-2.99) | 0.83 | 2.16(0.46-10.20) | 0.33 |
| LDL status (>100 mg/dl) | 3.00(1-9.01) | 0.052 | 3.46(1.03-11.59) | 0.045 |
| Statin use | 0.73(0.21-2.58) | 0.63 | 2.11(0.41-10.81) | 0.37 |

Figure 4 displays the impact of gender-affirming treatments on cardiovascular risk factors. Transgender individuals who had undergone surgical and/or hormonal treatment exhibited significantly higher odds of tobacco use (aOR, 6.67 [95% CI, 2.80-15.98]), statin need (aOR, 3.97 [95% CI, 1.14-13.81]), and alcohol use (aOR, 11.31 [95% CI, 1.34-95.53]). Our findings reveal a significant association between transgender status and specific cardiovascular risks factors, such as tobacco use and prediabetes/diabetes. Moreover, distinct risk profiles were identified for transgender males and females, with males experiencing higher rates of obesity and hyperlipidemia. Additionally, surgery and hormone treatment status were found to be associated with certain cardiovascular risk factors.

**Figure 4:**
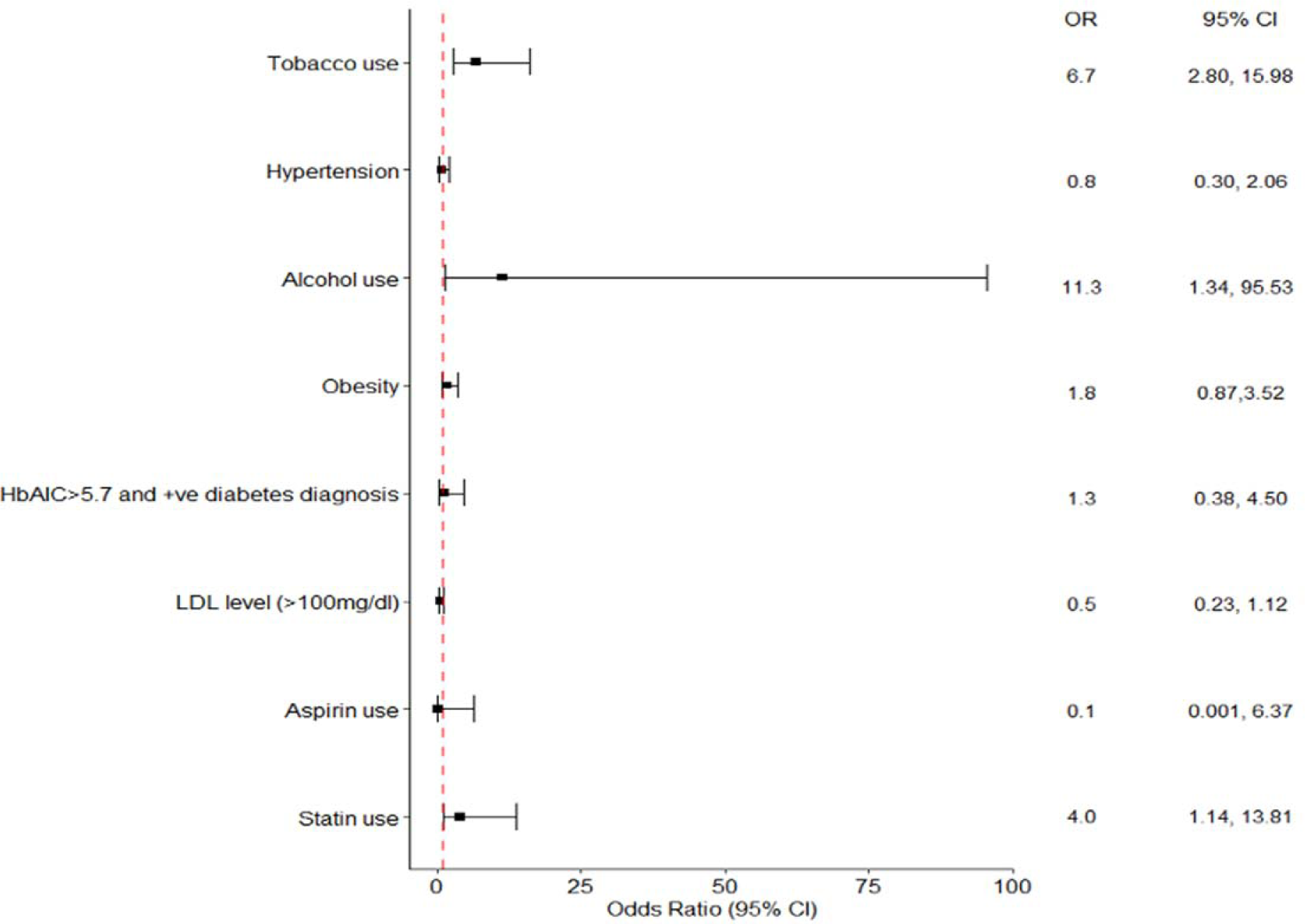
Adjusted multivariate logistic regression of cardiovascular risk factors in transgender patients who underwent surgery and/or hormone treatment vs. patients with no surgery and/or hormone treatment.

## Discussion

In this study, we comprehensively examine the prevalence and risk factors for smoking, obesity, hypertension, diabetes/insulin resistance, hyperlipidemia, and alcohol use within the transgender population, emphasizing the unique health challenges this group faces. It is particularly relevant to conduct such studies in socioeconomically disadvantaged communities like the Appalachian region, where health disparities are more pronounced and access to culturally competent care is limited.

The Appalachian region has long been associated with poverty, lower levels of education, and reduced access to healthcare services. These factors contribute to a heightened risk of chronic diseases and adverse health outcomes for the population residing in this area. The transgender community, already vulnerable due to existing healthcare access and utilization disparities, faces additional challenges in such settings. Understanding the specific health concerns of the transgender population in impoverished regions like Appalachia is crucial for developing targeted interventions and promoting health equity.

The following subsections discuss our results regarding tobacco use, obesity, hypertension, diabetes/insulin resistance, hyperlipidemia, and alcohol use. By addressing these risk factors collectively, healthcare providers can better understand the complex interplay of health challenges faced by transgender individuals and develop comprehensive strategies for prevention and treatment.

### Tobacco Use

Our study reveals a significantly higher prevalence of tobacco use among transgender individuals than cisgender individuals, with an adjusted odds ratio (aOR) of 6.64 (95% CI, 2.59-17.01) and higher odds of smoking in Transgender individuals that had either surgery or hormonal therapy (aOR) of 6.69 (95% CI, 2.80-15.98). This finding is consistent with prior research from the BRFSS data, which showed an increased prevalence of smoking in transgender patients (19.7%) compared to cisgender individuals (16.8%).^14^ The heightened prevalence of tobacco use within the transgender population may be attributed to the Minority Stress Theory (MST), which postulates that the internal stress resulting from gender non-affirmation, stigma, discrimination, and victimization can lead to unhealthy coping mechanisms, such as tobacco and drug use.^15–16^ Healthcare providers should be aware of the increased risk of tobacco use in transgender patients and provide appropriate support and counseling to help them quit smoking, potentially mitigating these risk factors, and promoting better cardiovascular health among transgender individuals.

### Alcohol Use

We report a significantly higher proportion of alcohol use among transgender individuals with a history of surgery or hormone therapy than those without (aOR 11.31 [95% CI, 1.34-95.53]), with a higher (aOR 7.07 [95% CI, 0.84-59.22]) proportion of alcohol use among transgender individuals (10.3%) compared to cisgender individuals (1.5%) without statistical significance. This aligns with prior research reporting a 36% prevalence of alcohol use disorder in the transgender population, compared to 8.5% in the general population.^17–18^ Transgender individuals have also been found to experience more negative alcohol-related consequences, such as blacking out or legal issues due to drinking.^19^ The higher prevalence of alcohol use in this population may be attributed to various factors, including coping mechanisms for dealing with discrimination and minority stress.^20^ It is crucial to address the psychosocial factors that may contribute to higher alcohol use rates among transgender individuals, such as minority stress, discrimination, and lack of social support. Culturally competent care and support systems should be established to help transgender patients cope with these challenges and reduce their reliance on alcohol as a coping mechanism.

### Obesity

We report a higher prevalence of obesity among transgender individuals than cisgender individuals, although not statistically significant (aOR 1.75 [95% CI, 0.87-3.52]). We observe similar findings in transgender individuals with a history of surgery or hormonal therapy. However, there is increased odds of obesity in the transgender male group compared to the transgender female group in our study (aOR, 13.39 [95% CI, 3.30-54.30]). This supports prior research suggesting that obesity rates are higher among transgender individuals, especially in female to male (FTM) individuals compared to male to female (MTF) individuals.^14^ The BRFSS study also reported increased obesity rates among transgender individuals compared to cisgender individuals.^14^ While the etiology of obesity in the transgender population is multifaceted, one prominent factor is gender-affirming hormone therapy, which can influence fat distribution and muscle mass.^21^ Psychological stressors and their impact on cortisol levels should be considered, as chronic stress can affect metabolism and weight gain. Other factors contributing to obesity in this population may include poor access to healthcare services, as seen in our study area.

### Diabetes/Insulin-Resistance

Our study reports a higher prevalence of diabetes and/or HbA1C > 5.7% among transgender individuals (16.5%) compared to cisgender individuals (4.4%). The adjusted odds ratio (aOR) for prediabetes/diabetes in the transgender population was 3.98 (95% CI, 1.05-15.15). This aligns with previous research reporting a five-fold increase in diabetes prevalence among transgender youth^22^ and increased insulin resistance among transgender men and women undergoing gender-affirming hormone therapy.^14–15^ Both physiological and psychosocial factors, such as hormone therapy and discrimination-induced chronic stress, may contribute to the increased risk.^16^ These findings highlight the importance of regular monitoring and tailored interventions for diabetes and insulin resistance management in transgender individuals. Future research should explore the specific mechanisms underlying these disparities and identify effective strategies for mitigating diabetes risk in this population.

### Hyperlipidemia

There were higher odds of hyperlipidemia and statin requirement among transgender individuals than cisgender individuals. The mean LDL levels were 105.8 mg/dl (±37.5) and 108.2 mg/dl (±31.3) among transgender and cisgender groups. However, the cisgender group had a higher proportion of high LDL levels (66.7% vs. 55.2%). Additionally, the transgender population had a higher statin use rate than the cisgender population (15.8% vs. 5.8%). Among the transgender population, transgender men had higher odds of hyperlipidemia than transgender women (aOR 3.46 [95% CI, 1.03-11.59]). These findings are consistent with previous research indicating that transgender men undergoing gender-affirming hormone therapy have a higher prevalence of dyslipidemia^18^ and significant changes in LDL-C, TG, and HDL-C after two years of hormone therapy.^23^ Healthcare providers should be vigilant in assessing lipid levels and managing cardiovascular risk factors in transgender individuals, particularly transgender men, and provide culturally competent care and education to address their unique needs and promote adherence to lipid-lowering therapies.

### Hypertension

Our findings indicate that the prevalence of hypertension was not significantly different between transgender and cisgender individuals (aOR 0.78 [95% CI, 0.30-2.06]). The incidence of hypertension was comparable between the transgender and general populations (16.7% and 15.9%, respectively). Despite multiple risk factors that could increase the development of hypertension among transgender individuals, the difference in prevalence between the cis and trans populations was not statistically significant (p=0.9056). This finding aligns with previous systematic reviews, which report minimal and clinically insignificant elevations in SBP readings among transgender patients.^21^ However, it is essential to note that some studies have observed differences in blood pressure among transgender women with higher testosterone levels.^4,24^ Although research in this area is limited, the contradictory reports on blood pressure changes in transgender women on GAHT warrant further investigation to understand the etiology of hypertension in this population.^25^

### Study Limitations

While our study provides novel insights into the cardiovascular risk factors among transgender individuals in rural Appalachia, and the association with hormone therapy, some limitations should be considered when interpreting the findings. Our study’s sample size may be relatively small, which could limit the generalizability of the findings to the broader transgender population. Larger, multi-center studies would provide more robust data and greater statistical power. Our study relied on self-reported data for certain variables, such as alcohol consumption, which can be subject to recall bias and social desirability bias, leading to the under or over-reporting of behaviors. Finally, there may be unmeasured confounding factors that could influence the associations observed between hormone therapy, risk factors, and cardiovascular outcomes, such as socioeconomic status and access to healthcare. Future research should address these limitations by employing longitudinal study designs, larger sample sizes, and more comprehensive assessments of hormone therapy regimens and potential confounding factors.

## Conclusion

We shed additional light on the increased prevalence of tobacco use, obesity, hypertension, diabetes/insulin resistance, hyperlipidemia, and alcohol use among the transgender population compared to their cisgender counterparts in rural Appalachia. These findings emphasize the importance of developing targeted interventions and raising awareness among healthcare providers to address the specific health needs of this population. By acknowledging and addressing these disparities, we can work towards improving the overall health and well-being of transgender individuals and reducing their cardiovascular risk.

## Disclosures

Dr. Rodriguez is an iTHRIV Scholar. The iTHRIV Scholars Program is supported in part by the National Center for Advancing Translational Sciences of the National Institutes of Health under Award Numbers UL1TR003015 and KL2TR003016.

All other authors have reported that they have no relationships relevant to the contents of this paper to disclose.

## Data Availability

The data that support the findings of this study are available from the corresponding author, EE, upon reasonable request.

---

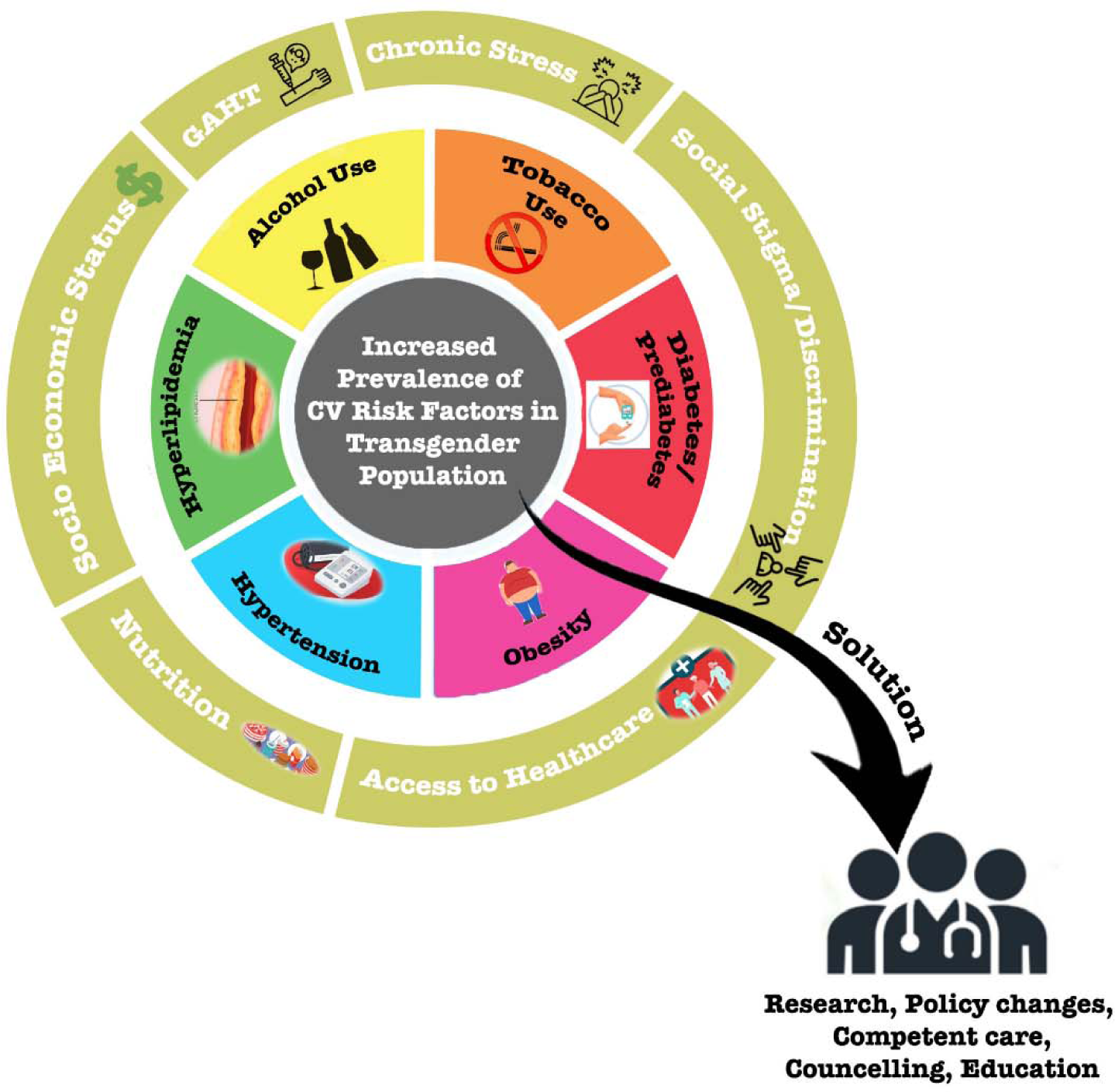
Central Illustration: Cardiovascular risk factors among the Appalachian transgender population and proposed solutions.

## Non-standard Abbreviations and Acronyms

CVD: Cardiovascular diseases TGD Transgender
GAHT: Gender-affirming hormone therapy BMI Body mass index
CAD: Coronary artery disease LDL Low-density lipoprotein HbA1C Glycated hemoglobin DM Diabetes Mellitus
FTM: Female to male MTF Male to female
HIV: Human Immunodeficiency Virus

## Notes

### Funding Statement

No funding was needed or received.

### Author Declarations

The Marshall University Institutional Review Boards and Ethics Review Board approved the study.

## References

1. Roth GA, Mensah GA, Johnson CO, Addolorato G, Ammirati E, Baddour LM, et al. Global Burden of Cardiovascular Diseases and Risk Factors, 1990-2019: Update From the GBD 2019 Study. J Am Coll Cardiol. 2020 Dec 22;76(25):2982-3021.

2. American Psychological Association. Guidelines for psychological practice with transgender and gender nonconforming people. Am Psychol. 2015 Dec;70(9):832–64.

3. Human Rights Campaign Foundation. (2021). We are Here: Understanding the Size of the LGBTQ+ Community. Human Rights Campaign Foundation. https://hrc-prod-requests.s3-us-west-2.amazonaws.com/We-Are-Here-120821.pdf

4. Wierckx K, Elaut E, Declercq E, Heylens G, De Cuypere G, Taes Y, et al. Prevalence of cardiovascular disease and cancer during cross-sex hormone therapy in a large cohort of trans persons: a case-control study. Eur J Endocrinol. 2013 Oct;169(4):471–8.

5. Maraka S, Singh Ospina N, Rodriguez-Gutierrez R, Davidge-Pitts CJ, Nippoldt TB, Prokop LJ, et al. Sex Steroids and Cardiovascular Outcomes in Transgender Individuals: A Systematic Review and Meta-Analysis. J Clin Endocrinol Metab. 2017 Nov 1;102(11):3914–23.

6. How Many Adults and Youth Identify as Transgender in the United States? Williams Institute. https://williamsinstitute.law.ucla.edu/publications/trans-adults-united-states/. Accessed on April 25, 2023.

7. Mamudu HM, Paul TK, Wang L, Veeranki SP, Panchal HB, Alamian A, Budoff M. Association Between Multiple Modifiable Risk Factors of Cardiovascular Disease and Hypertension among Asymptomatic Patients in Central Appalachia. South Med J. 2017 Feb;110(2):90–96.

8. Coleman E, Bockting W, Botzer M, Cohen-Kettenis P, DeCuypere G, Feldman J, et al. Standards of Care for the Health of Transsexual, Transgender, and Gender-Nonconforming People, Version 7. International Journal of Transgenderism. 2012 Aug 1;13(4):165–232.

9. Rider GN, McMorris BJ, Gower AL, Coleman E, Eisenberg ME. Health and Care Utilization of Transgender and Gender Nonconforming Youth: A Population-Based Study. Pediatrics. 2018 Mar;141(3):e20171683.

10. Conron KJ, Scott G, Stowell GS, Landers SJ. Transgender Health in Massachusetts: Results From a Household Probability Sample of Adults. Am J Public Health. 2012 Jan;102(1):118–22.

11. Meerwijk EL, Sevelius JM. Transgender Population Size in the United States: a Meta-Regression of Population-Based Probability Samples. Am J Public Health. 2017 Feb;107(2):e1–8.

12. Streed CG Jr, Beach LB, Caceres BA, Dowshen NL, Moreau KL, Mukherjee M, et al. Assessing and Addressing Cardiovascular Health in People Who Are Transgender and Gender Diverse: A Scientific Statement From the American Heart Association. Circulation. 2021 Aug 10;144(6):e136–e148.

13. Denby KJ, Cho L, Toljan K, Patil M, Ferrando CA. Assessment of Cardiovascular Risk in Transgender Patients Presenting for Gender-Affirming Care. Am J Med. 2021 Aug;134(8):1002–8.

14. Nokoff NJ, Scarbro S, Juarez-Colunga E, Moreau KL, Kempe A. Health and Cardiometabolic Disease in Transgender Adults in the United States: Behavioral Risk Factor Surveillance System 2015. J Endocr Soc. 2018 Mar 5;2(4):349–60.

15. Streed CG, Beach LB, Caceres BA, Dowshen NL, Moreau KL, Mukherjee M, et al. Assessing and Addressing Cardiovascular Health in People Who Are Transgender and Gender Diverse: A Scientific Statement From the American Heart Association. Circulation. 2021 Aug 10;144(6):e136–48.

16. Poteat TC, Divsalar S, Streed CG, Feldman JL, Bockting WO, Meyer IH. Cardiovascular Disease in a Population-Based Sample of Transgender and Cisgender Adults. Am J Prev Med. 2021 Dec;61(6):804–11.

17. Maru J, Millington K, Carswell J. Greater Than Expected Prevalence of Type 1 Diabetes Mellitus Found in an Urban Gender Program. Transgend Health. 2021 Feb;6(1):57–60.

18. Moreira Allgayer RMC, Gustavo da SB, Ramos RB, Filho RSM, Mara Spritzer P. RF34 | PMON276 The Effect of Gender Affirming Hormone Therapy (GAHT) on Non-Conventional Cardiovascular Risk Markers in Transgender Men: A Systematic Review. J Endocr Soc. 2022 Nov 1;6(Suppl 1):A717–8.

19. Hughto JMW, Pletta D, Gordon L, Cahill S, Mimiaga MJ, Reisner SL. Negative Transgender-Related Media Messages Are Associated with Adverse Mental Health Outcomes in a Multistate Study of Transgender Adults. LGBT Health. 2021 Jan 1;8(1):32–41.

20. Aranda G, Halperin I, Gomez-Gil E, Hanzu FA, Seguí N, Guillamon A, et al. Cardiovascular Risk Associated With Gender Affirming Hormone Therapy in Transgender Population. Frontiers in Endocrinology [Internet]. 2021 [cited 2023 Apr 25];12. Available from: https://www.frontiersin.org/articles/10.3389/fendo.2021.718200

21. Seal LJ. Cardiovascular disease in transgendered people: A review of the literature and discussion of risk. JRSM Cardiovasc Dis. 2019;8:2048004019880745.

22. Chobanian AV, Bakris GL, Black HR, Cushman WC, Green LA, Izzo JL, et al. Seventh report of the Joint National Committee on Prevention, Detection, Evaluation, and Treatment of High Blood Pressure. Hypertension. 2003 Dec;42(6):1206–52.

23. Abosi-Appeadu K, Maertelaere ASD, Shadid S, Defreyne J, T G. Does gender-affirming hormone therapy alter insulin resistance in transgender persons? In: Endocrine Abstracts [Internet]. Bioscientifica; 2018 [cited 2023 Apr 25]. Available from: https://www.endocrine-abstracts.org/ea/0056/ea0056p366

24. Chan Swe N, Ahmed S, Eid M, Poretsky L, Gianos E, Cusano NE. The effects of gender-affirming hormone therapy on cardiovascular and skeletal health: A literature review. Metabol Open. 2022 Mar;13:100173.

25. Wierckx K, Van Caenegem E, Schreiner T, Haraldsen I, Fisher AD, Toye K, et al. Cross-sex hormone therapy in trans persons is safe and effective at short-time follow-up: results from the European network for the investigation of gender incongruence. J Sex Med. 2014 Aug;11(8):1999–2011.

